# Design and interpretation of eQTL-GWAS colocalisation studies: lessons from a large-scale evaluation

**DOI:** 10.1101/2025.11.20.25340664

**Authors:** Guillermo Reales, Jeffrey M. Pullin, Ichcha Manipur, Elena Vigorito, Chris Wallace

## Abstract

Colocalisation analysis is extensively applied across diverse GWAS and molecular QTL datasets to identify candidate causal genes. We systematically characterised large-scale colocalisation results across eQTL studies varying in cellular granularity and sample size, with the goal of providing design and interpretation recommendations. We found 34-50% of GWAS hits colocalised, and were more likely to colocalise if they were located nearer genes and had a more common lead variant. We also found over 50% of colocalisations were found in only one cell type. This led to an inherent trade-off: while high granularity studies tended to have smaller sample sizes and lower eQTL discovery, each eQTL from these high-granularity datasets were more likely to colocalise, reflecting cell-type specificity. On the other hand, lower granularity studies achieved larger sample size and higher eQTL discovery, leading to detection of the greatest total number of colocalisations, particularly for lower frequency GWAS lead variants. This suggests large, high granularity studies will be needed to identify remaining colocalisations.

Of the peaks that colocalised, 37-47% did so with multiple genes, suggesting coregulation of the GWAS trait, horizontal pleiotropy, or false positives. However, sensitivity analyses indicated that even extremely stringent significance thresholds did not substantially reduce multi-gene colocalisations, arguing against widespread false discovery. Integration of enhancer–promoter interaction data provided evidence for coregulation among multi-colocalising eGenes. While disentangling causality from horizontal pleiotropy will ultimately require experimental perturbation, triangulation using different sources of observational data is likely to be necessary, provided careful consideration is taken to identify biases and missing data that may influence gene prioritisation.

## Introduction

Genome-wide association studies (GWAS) have identified hundreds of thousands of associations between genetic loci and traits of interest. However, mapping the path from genotype to phenotype remains challenging, due to the non-coding nature of more than 90% of the implicated variants, the small effect size of the vast majority of associations, the complexity posed by linkage disequilibrium and the fact that some loci contain multiple causal variants. One approach to elucidate the regulatory mechanisms is to integrate summary data from GWAS and expression QTLs (eQTLs) using colocalisation to nominate the likely causal genes for each GWAS signal. This approach aims to determine whether the GWAS trait and eQTL share a causal variant in a region, with the implicit conclusion that this is likely to indicate the eGeneQTL is on the causal pathway for disease.

Despite extensive efforts to link eQTLs from a wide range of tissue types to a diverse set of GWAS traits, most GWAS loci remain unexplained by currently catalogued eQTLs. For example, an analysis using GTEx expression data on 49 tissues and covering 5,385 GWAS loci, estimated the proportion of colocalising loci with at least one gene to be 43%^1^ or 40%^2^, with a median of ∼20% loci per trait. This has led to the notion of the so-called “colocalisation gap”: the fraction of GWAS signals with gene regulation potential that cannot be linked to a target gene by colocalisation.

Several hypotheses have been proposed to account for the missing colocalisation. One obvious explanation is that non-coding variants may have regulatory functions beyond gene expression. However, even when eQTLs, splicing-QTLs and protein-QTLs from 92 cell types were colocalised with 70,364 trait associated loci, the proportion of colocalised loci only increased to 50.6%^3^. Another possibility is that eQTL studies are not well powered for rarer variants, which are captured in GWAS due to their larger sample size. It has also been suggested that eQTLs and GWAS variants have other systematic differences, with eQTLs particularly enriched in promoters and typically situated much closer to genes, in addition to differences in functional annotations^4^. While this analysis holds for steady-state canonical eQTLs, other modalities differ. For example, enhancer RNAs tend to have more distal eQTLs compared to canonical genes, a profile more similar to GWAS hits^5^. Indeed, including eRNAs along with canonical gene eQTLs led to a 63% increase in the proportion of GWAS signals showing colocalisation. Additionally, we know that different sources for the eQTL study vary in how informative they are for any given GWAS. For example, eQTLs from patient samples are better able to explain disease associated GWAS signals than from healthy control samples^6,7^, just as samples from the disease site are more informative than samples of the same cell type from elsewhere in the body (eg immune cells from the colon rather than peripheral blood in inflammatory bowel disease, IBD)^8^. As well as sample source, the granularity of expression studies can vary, from bulk sequencing of mixed cells, through sorted cell populations to single cells (with reduced sample size tending to associate with increased granularity). This is important because many signals have shown to be context dependent, so depth and breadth are required to detect cell state specific signals^9^, or ancestry dependent signals.

For a GWAS practitioner wanting to use molecular eQTL studies to aid interpretation of their new GWAS results, or for an investigator deciding whether to invest in a new eQTL study, it can be unclear which source of eQTLs are most likely to yield interpretable results. We set out to explore two massively parallel colocalisation analyses, one global using results from Open Targets Genetics, and a second focused on immune-mediated diseases (IMD) and immune cells, in order to estimate the colocalisation gap and determine the most informative source of eQTL studies. While most signals colocalised with only one gene, we also observed colocalisation with two or more genes, similar to previous reports^2,3^. This hinders interpretation with regards to the causal pathway from a variant to phenotype. Which is/are the causal(s) gene(s) and which is the relevant cell type(s)? We integrated promoter enhancer interaction data^10^ with colocalisation results to investigate whether these multiple colocalisations were likely to represent false positives or co-regulation of neighbouring genes.

## Results

### 50% of GWAS signals in the open targets genetics (OTG) resource colocalise with at least one gene

We began by considering the ∼ 1.3x10^6^ colocalisation tests available in Open Targets Genetics (OTG). These consist of pairwise comparisons of up to 126 eQTL datasets with 18,126 eGenes from 100 tissues or cell types and 4,687 GWAS datasets covering 24 categories of disease or traits, leading to 159,585 fine mapped GWAS peaks. They represent the largest available public massively parallel set of colocalisation analyses to our knowledge.

The number of coloc-positive tests is a function of the threshold, α, on the posterior probability of colocalisation, PP(H4), used to call a positive test (Fig. 1A). If PP(H4) is well calibrated, the expected false discovery rate (FDR) amongst all tests with PP(H4) > α is simply the mean of (1-PP(H4)) amongst that set of tests. We chose α as the smallest value for which the estimated FDR < 0.05 (Fig.1B), giving a value of α=0.86. At this threshold, we found that overall 50.5% (95% confidence interval: 50.3%-50.8%) of loci colocalised with at least one gene, with a median of 33% per trait (Fig. 1C). Throughout the manuscript, “significant colocalisation” refers to results meeting this 5% FDR criterion.

**Figure 1.**
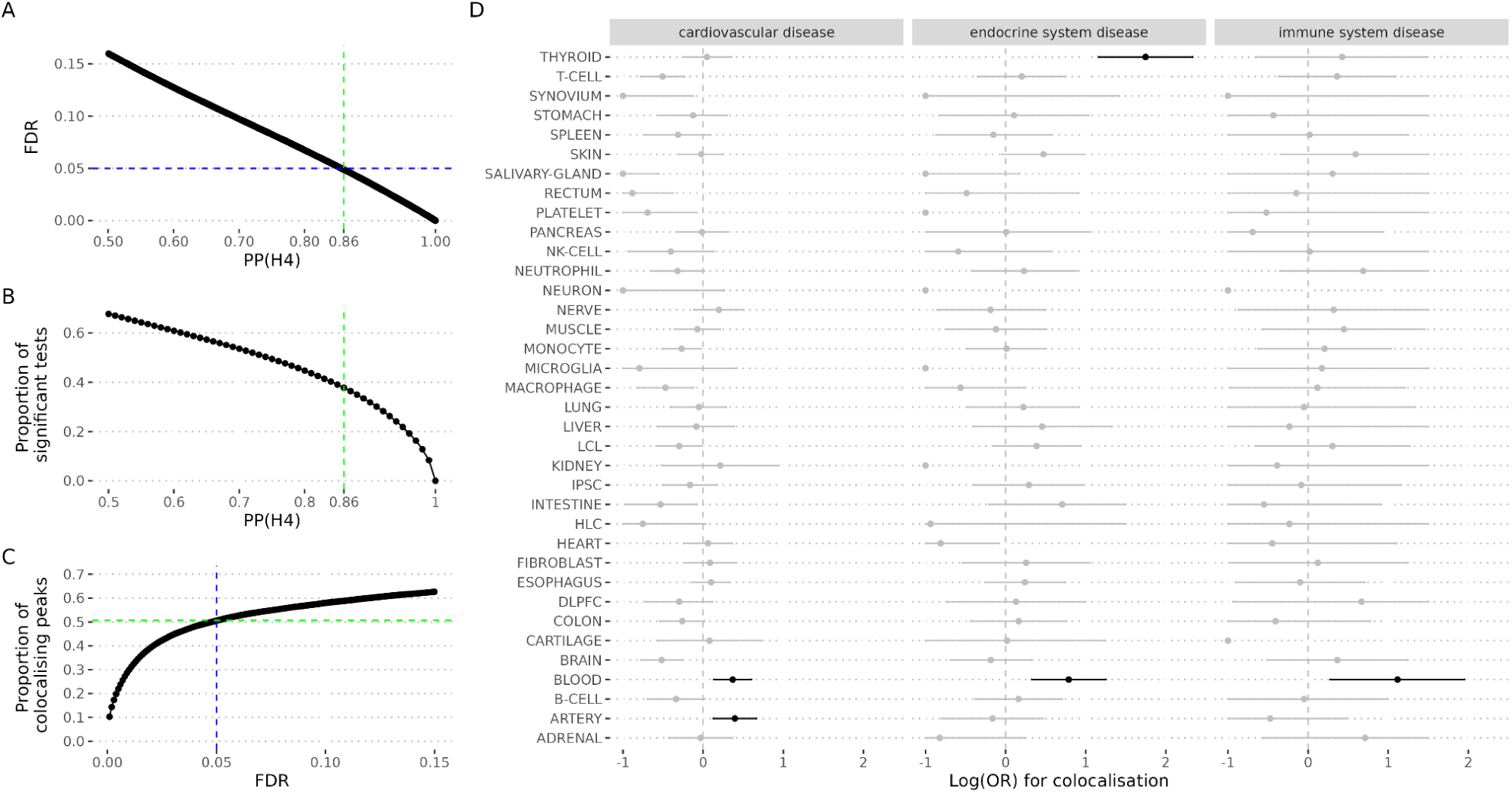
OTG colocalisation gap. **A.** Estimated false discovery rate (FDR) (y-axis) as a function of the posterior probability for colocalisation (PP(H4)) (x-axis). **B.** Proportion of coloc significant tests (y-axis) as a function of the posterior probability of colocalisation (PP(H4)). **C**. Proportion of peaks with evidence of colocalisation. In A,B and C, the vertical and horizontal lines indicate the thresholds selected from downstream analysis that corresponds to 5% FDR. **D**. Log(OR) for colocalisation relative to adipose tissue and 95% adjusted credible intervals by the number of comparisons (x-axis) in different cell types or tissues (y-axis) for three exemplar disease traits. Black symbols indicate significant enrichment. Effect sizes were truncated at -1.

### Colocalisation is more likely in disease-relevant cell types

We anticipated that colocalisation rates might vary between tissues. To explore this hypothesis, we selected all colocalisation tests for GWAS from three of the OTG categories: “immune system disease” (3,144 tests), “endocrine system disease” (8,331 tests) and “cardiovascular disease” (30,111 tests) for which we could identify likely causal tissues or cell types. All showed enrichment in biologically relevant eQTL datasets: immune system disease in blood, cardiovascular disease in artery and blood, and endocrine system disease in thyroid and blood (Fig. 1D). This analysis supports another common approach to colocalisation analysis, focused on *a priori* likely causal cell types. To investigate this design, we considered a more specific set of colocalisation analyses, focused on immune-mediated disease (IMD) GWAS and their biologically relevant immune cell eQTLs. Immune cell eQTLs offer an opportunity to explore a rich diversity of designs, and we assembled a catalog of 12 eQTL studies (five not available in OTG) covering a total of 101 eQTL experiments from peripheral blood cells (Supp. Table 1). This allowed us to consider the importance of the eQTL granularity - whether mixed cell populations with their attendant larger sample sizes could be more useful than smaller, sorted or single cell populations: our catalog combined a very well powered whole blood eQTL studies (eQTLGen^11^ with sample size N >30K), with sorted and single cell eQTL studies on a wide range of cell types (>20 cell types for ImmunexUT^12^ or OneK1K^13^, albeit with N under 1000). While most studies were of European ancestry, ImmunexUT participants were Japanese while other studies combined European and Asians^14^ or Africans^15^. We also addressed whether samples from patients may be more informative than those from healthy controls^7^ by including eQTL studies combining IMD donors and controls or stimulated cells (Supp. Table 1 and Supp Fig. 1).

We colocalised eQTLs from each of these datasets with 14 exemplar IMD GWAS covering 10 distinct IMDs (Supp. Table 2). Overall, we performed 153,656 colocalisation tests, related to 832 GWAS peaks. We also included six non-IMD diseases (Supp Table 2) for comparison for which we conducted 65,272 additional coloc tests. We again applied a 5% FDR to define significant colocalisations, which corresponded to α=0.88 (Supp. Fig. 2A-B). Our results (Supp. Tables 3 and 4) were similar overall to those from the broader OTG study, although we found that only 34% of GWAS peaks colocalised with at least one eQTL (Figure 2A), possibly due to the narrower cell type coverage in this focused design. Otherwise, colocalisation rates were not significantly related to the power of the GWAS study (Fig. 2A,B,H) and were higher for IMDs (when the cell types used in the eQTL studies play a role in the disease process) than compared to a set of non-IMD GWAS (Supp. Fig. 2C), with an odds ratio for IMD colocalisation of 1.6 (1.4-2.3), relative to non-IMDs. We found that protein coding genes alone were able to explain ∼90% of the GWAS peaks with a colocalisation, while the remaining 10% could only be explained by non-coding RNAs.

**Figure 2.**
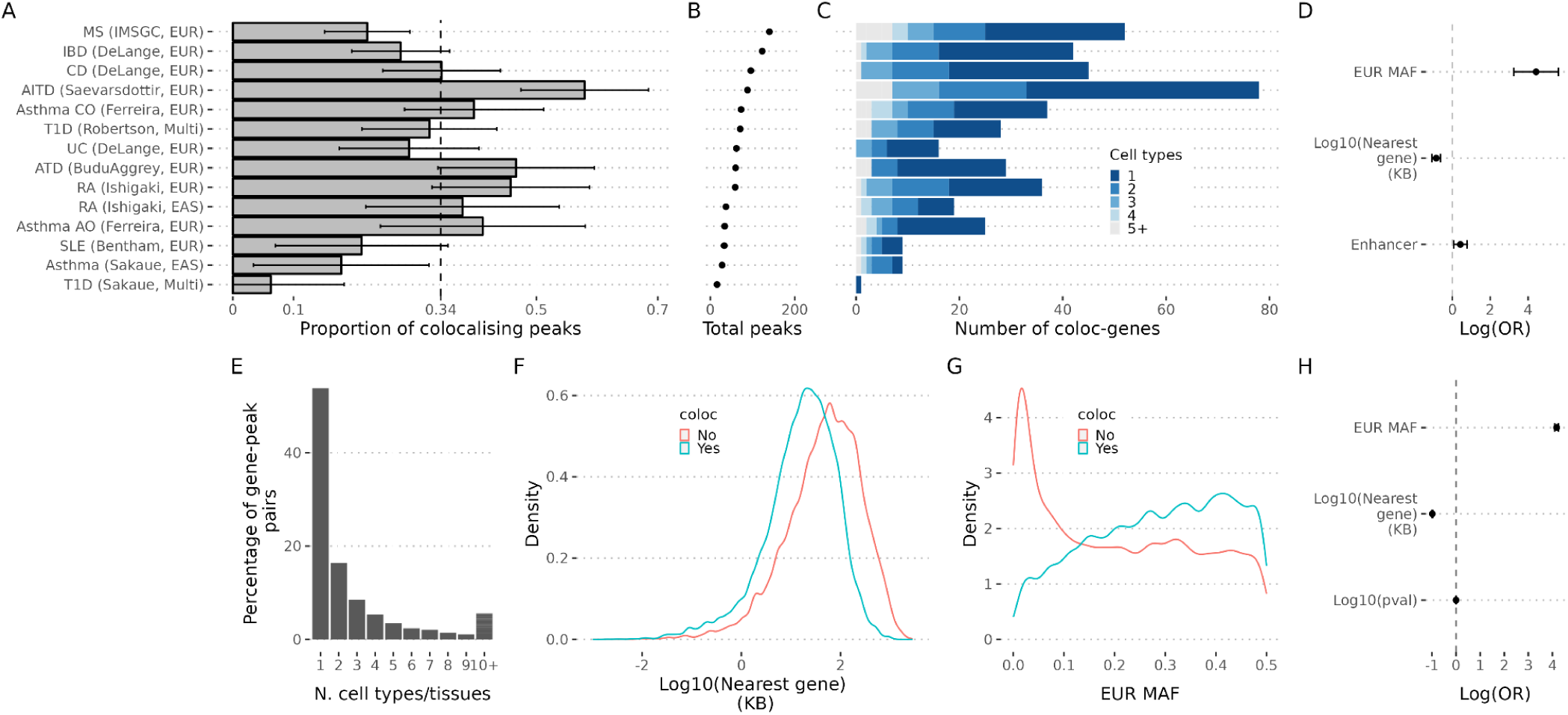
Colocalisation gap by IMD and characteristics of GWAS lead variants stratified by colocalisation status . A-D correspond to the analysis of IMDs. **A**. Proportion of GWAS loci with at least one significant coloc test (bar plot) for each IMD. The dashed vertical line represents the average proportion, 0.34 [0.30 - 0.36]. **B**. The dot plot shows the total number of GWAS loci by IMD. **C.** Cell specificity of colocalising genes. eQTL studies were aggregated by nominal cell types (CD4 T, CD8T, double negative T, gamma delta T, Monocyte, Neutrophil, B, DC, NK, HSPC, MAIT, Platelet). For each gene colocalising with a peak, the number of cell types that the colocalisation was detected was recorded. The plot shows the number of colocalising gene-pairs stratified by the number of cell types detected (x-axis) by IMD (y-axis). **D**. The plot shows the log odds ratio for colocalisation in the IMD study when the GWAS lead variant from an IMD overlaps an active enhancer in immune cells (Enhancer), for the allele frequency variance of the GWAS lead variant or for the log distance in kilo base pairs in base 10 between the GWAS lead variant and the nearest gene (Log10(distance to nearest gene (KB)). **E-H corresponds to OTG**.**E**. For each gene clocalisaing with a peak we counted the number of cell types or tissues that a significant colocalisation was observed. The plot shows the proportion of gene-peak pairs detected in the indicated number of cell types or tissues (x-axis). The plot was truncated at 10 cell types or tissues. **F.** Density plot for log distance in kilo base pairs in base 10 between the GWAS lead variant and the nearest gene stratified by colocalisation status. **G**. Same as F but for the European minor allele frequency. **H.** Log odds ratio for colocalisation in the OTG study for European minor allele frequency, distance to nearest gene and log in base 10 for the trait association.

We then compared the properties of fine mapped lead GWAS variants (as proxies for the causal variants) with or without evidence of colocalisation. Mostafavi et al^4^ recently explored the absence of colocalisation identified for many GWAS signals, describing significant differences in the patterns of GWAS and eQTL association signals. In agreement with this, we found that lead variants of GWAS signals with significant colocalisation results tended to be closer to a gene transcription start site (TSS) than those without colocalisation. This result was consistent in the IMD focussed analysis and in the global OTG (Fig. 2D,F,H). Moreover, we also saw that lead GWAS variants with evidence of colocalisation tended to have higher minor allele frequency in both analyses (Fig. 2D,G,H), suggesting a lack of power to detect potential eQTLs for lower-frequency variants in current eQTL studies. In addition, IMD GWAS lead variants tended to be within predicted active enhancers for PBMCs (Fig. 2D). The nearest gene to a GWAS peak is usually considered the most likely causal gene, supported by our results in IMDs which linked 67% of significant colocalisations to the closest gene (Supp. Fig. 2D).

### Colocalisation enrichment in more granular designs though studies still underpowered

We exploited the diversity of our IMD-focused panel of colocalisation tests to assess how eQTL study design impacted colocalisation discovery. We first tested for enrichment in the proportion of colocalisation significant tests within nominal immune cell types, that is, the chance of colocalisation conditional on detecting an eQTL. Relative to whole blood, CD8 and CD4 T cells, B cells and dendritic cells showed significant enrichment, as did single cell eQTL datasets relative to sorted cells, and eQTL experiments with a mixture of patients and control samples, relative to control samples only, (Fig 3A-B). In contrast, the sample of stimulated cells we considered did not show enrichment (Fig. 3B). While whole blood is not enriched in discoveries, this is compensated by the large sample size which leads to increased eQTL discovery: more colocalising genes are found in eQTLGen than any other dataset (Supp. Fig. 3), and eQTLGen is often the only source of colocalisations for GWAS peaks with rarer variants (Fig 3C), suggesting the lack of power associated with smaller sample sizes for more sophisticated designs is an important issue^16^. It is important to note, however, that using only eQTLGen or even all blood samples would not be sufficient: 48% of GWAS peaks that showed colocalisation only did so using sorted or single cell eQTL studies, compared to 15% that showed colocalisation only in whole blood.

**Figure 3.**
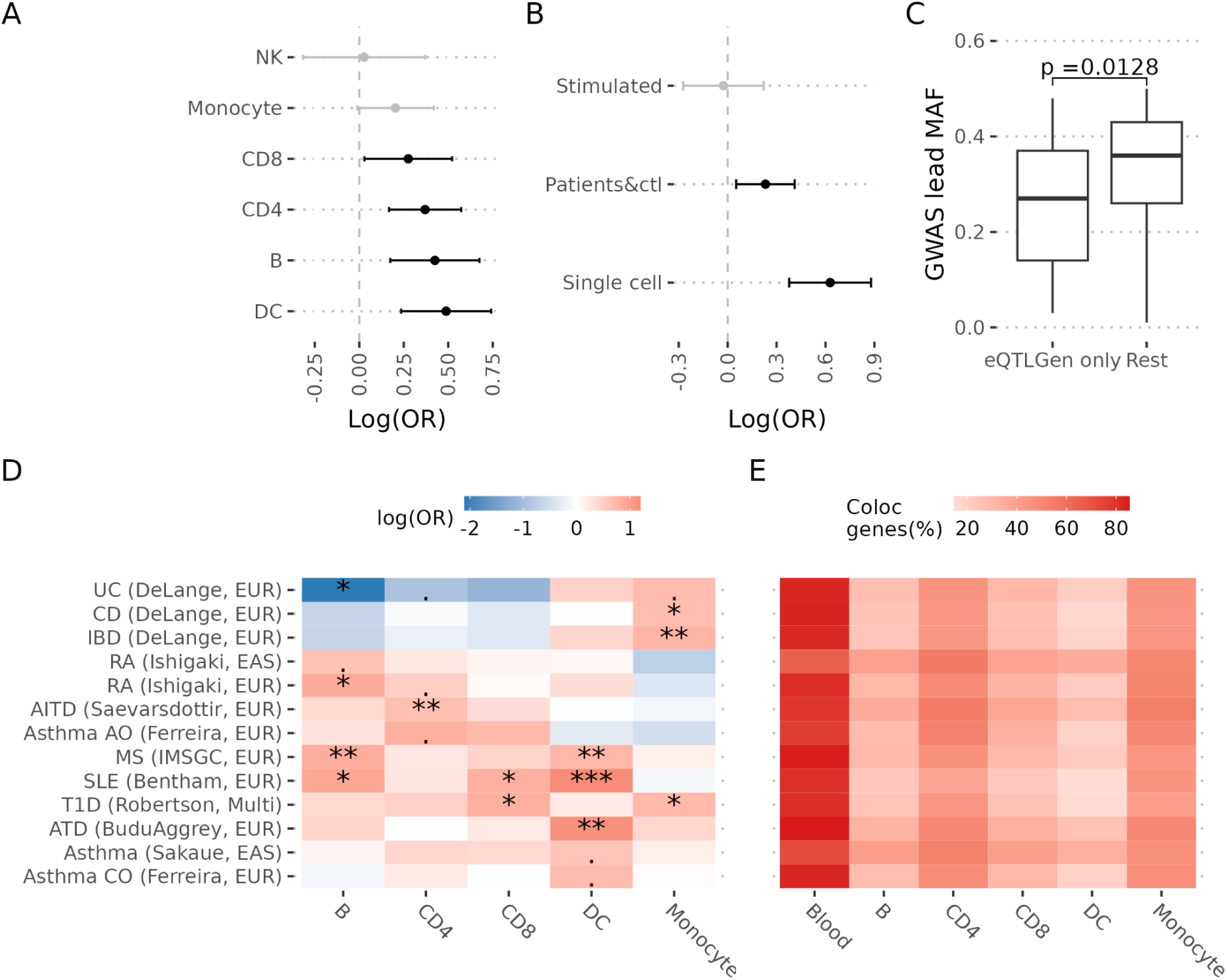
Trade-off of different eQTL designs. This analysis was performed for IMDs. **A**. Colocalisation odds ratio (x-axis) for the indicated cell types (y-axis) relative to whole blood. **B.** Colocalisation odds ratio (x-axis) for single cell eQTL studies relative to sorted cell, stimulated relative to steady state and mixture of patients and controls (Patients&ctl) relative to controls. **C**. Allele frequency distribution for the GWAS SNP (y-axis) stratified by those which only showed significant coloc tests in eQTLGen vs those detected in any other eQTL experiment (rest). **D.** Heatmap showing the enrichment for colocalisation for the indicated cell types (x-axis) relative to whole blood stratified by IMD (y-axis). The symbols indicate significance level (. 0.05 ⪯ p < 0.1, * 0.01 ⪯ p < 0.05, ** 0.001 ⪯ p < 0.01, *** p < 0.001). **E**. Heatmap showing the proportion of coloc significant genes identified by each of the indicated cell types (x-axis) for the IMDs in the y-axis.

We also saw considerable evidence of cell-type specificity. In OTG, with a broad range of tissues, we found that 53% of the peaks with a significant colocalisation test with a particular gene were detected in only one tissue/cell type (Fig. 2E). In our IMD study, we found that 55% of colocalising genes were detected in one nominal cell type only (CD4 T, CD8 T, double negative T, gamma delta T, MAIT, Monocyte, Neutrophil, B, DC, NK, HSPC, Platelet), with a rapid decrease in the number of genes colocalising in multiple cell types, a pattern that was shared across IMDs (Fig. 2C). Additionally, we found specific patterns of colocalisation enrichment across IMDs, recapitulating known biology. For example, Crohn’s disease and inflammatory bowel disease colocalisation signals were enriched in monocytes, while systemic lupus erythematosus colocalisation signals were enriched in B cells and dendritic cells (Fig. 3D). However, when we looked at the proportion of colocalisations by cell type/tissue in each IMD, whole blood was the predominant contributor across the board, reflecting again the increased power of eQTLGen (Fig. 3E).

### Genes colocalising with the same GWAS peak show evidence for co-regulation

We found that ∼47% of GWAS peaks with significant colocalisation in OTG and 37% in the IMD analysis displayed this colocalisation with two or more genes, though the number of peaks with more than two colocalising genes rapidly decreases (Fig. 4A). Among the peaks with multiple colocalising genes in the IMD analysis, approximately half (60/118) colocalised with protein-coding genes only, 47% (55/118) colocalised with both protein-coding and non-coding RNA genes, and the remaining ∼3% (3/118) colocalised exclusively with non-coding RNA genes.

**Figure 4.**
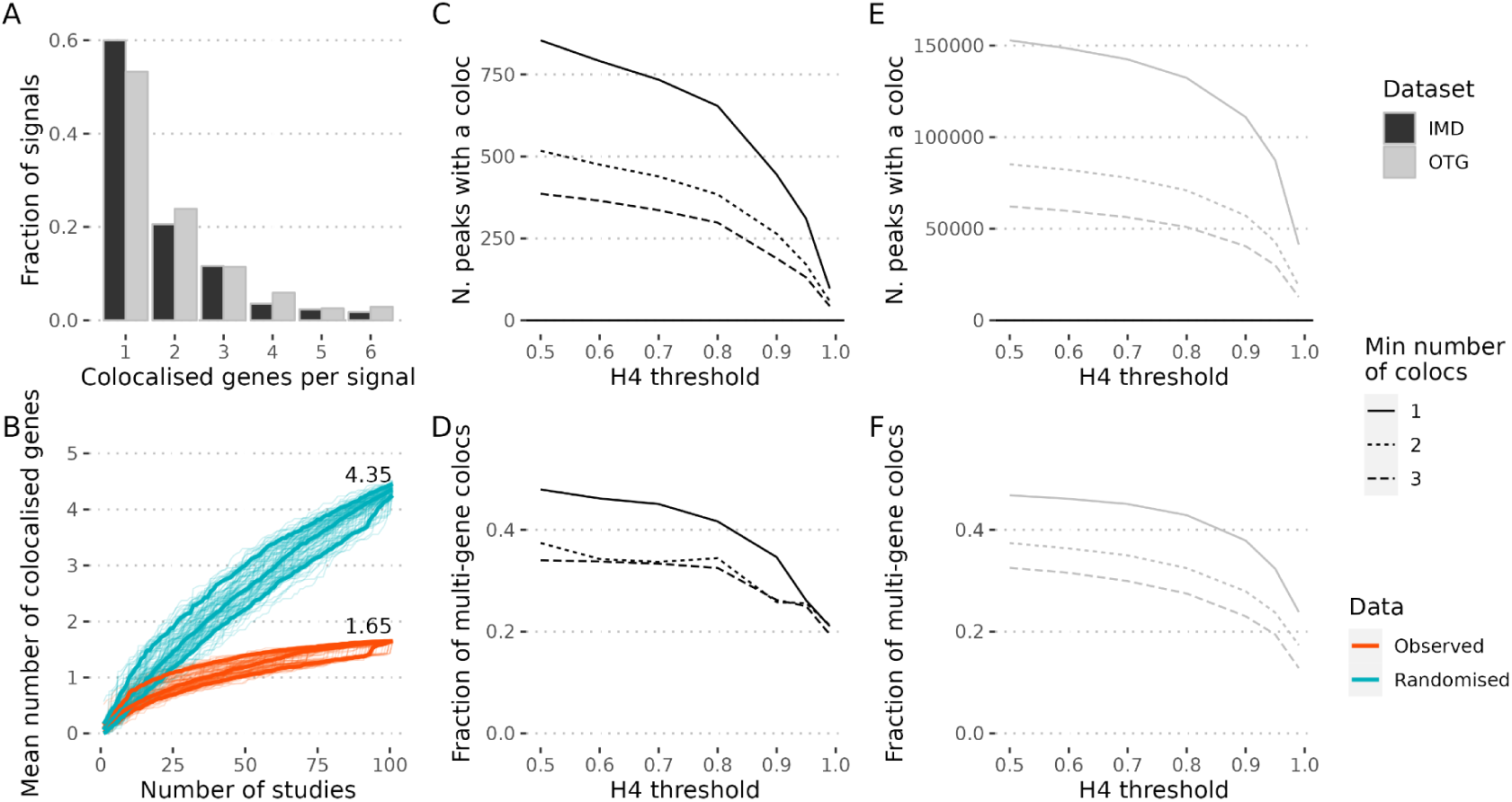
Assessing false positive colocalisation calls within multi-gene coloc peaks. **A.** Distribution of the proportion of colocalised genes per peak in the IMD (black bars) or OTG (grey bars) analysis. **B.** Cumulative mean of the number of colocalised genes after adding each study. The order of the 101 eQTL studies was randomised 100 times. In each iteration the cumulative mean of the colocalised genes across all peaks was calculated after the addition of each study, either with the observed data (orange) or on previously randomised PP(H4) within each peak (blue). Each iteration is represented by a fine line. The thicker lines correspond to the 10, 50 and 90% quantiles. The numbers in the plot correspond to the median of the iteration means after the addition of all the studies. **C,E.** At each of the indicated thresholds for the posterior probability of H4 (x-axis), the number of peaks with at least 1 (solid line), 2 (dotted line) or 3 (dashed line) colocalising genes is represented is shown for the IMD analysis (C) or OTG (E), respectively. **D,F.** Similar to C,E except that the fraction of multi-gene colocalisations across peaks is shown for the IMD analysis (D) or OTG (F), respectively.

One possibility to explain the number of peaks with multiple colocalising genes is that coloc is failing to control the false discovery rate, so that with increasing numbers of eQTL datasets we find more colocs, and that the multiple colocalisations simply represent false discoveries. We compared the observed mean number of colocalising genes per GWAS peak to that expected under a model of random colocalisation using the IMD analysis, and found considerable concentration of colocalisation signals within genes in the observed data (Fig. 4B). We also examined how the number of GWAS peaks with multiple colocalising genes changed as a function of increasingly stringent thresholds for calling colocalisation (Fig. 4C,E). Even when increasing stringency to the point of reducing the number of GWAS peaks with colocalisations from 854 to 97 in the IMD analysis or from 152K to 41K in OTG, the fraction with multiple colocalising genes remained above 20% for IMD or 15% for OTG (Fig. 4D, F). While we cannot rule out false colocalisations, and indeed expect 5% of our calls will be false given we targeted 5% FDR, these findings suggest that false positives alone cannot explain the multi-gene colocalisations.

An alternative is that they reflect shared regulation, a hypothesis supported by evidence for proximal genes to be co-expressed and share regulatory elements^17–19^ . After excluding blood (a mixed tissue), we identified 549 non-overlapping GWAS peaks where at least two genes were tested for colocalisation (see Methods). At each peak we considered all pairs of tests formed from distinct genes. We found that both tests were more likely to be significant colocalisations if they were from the same cell type compared to different cell types (odds ratio=1.2, p=0.004). We also found that pairs of genes colocalising with the same peak tended to show higher correlation in expression, and to be closer to each other, relative to pairs of colocalising and non-colocalising eGenes tested at the same GWAS signal Fig. 5A-B).

**Figure 5.**
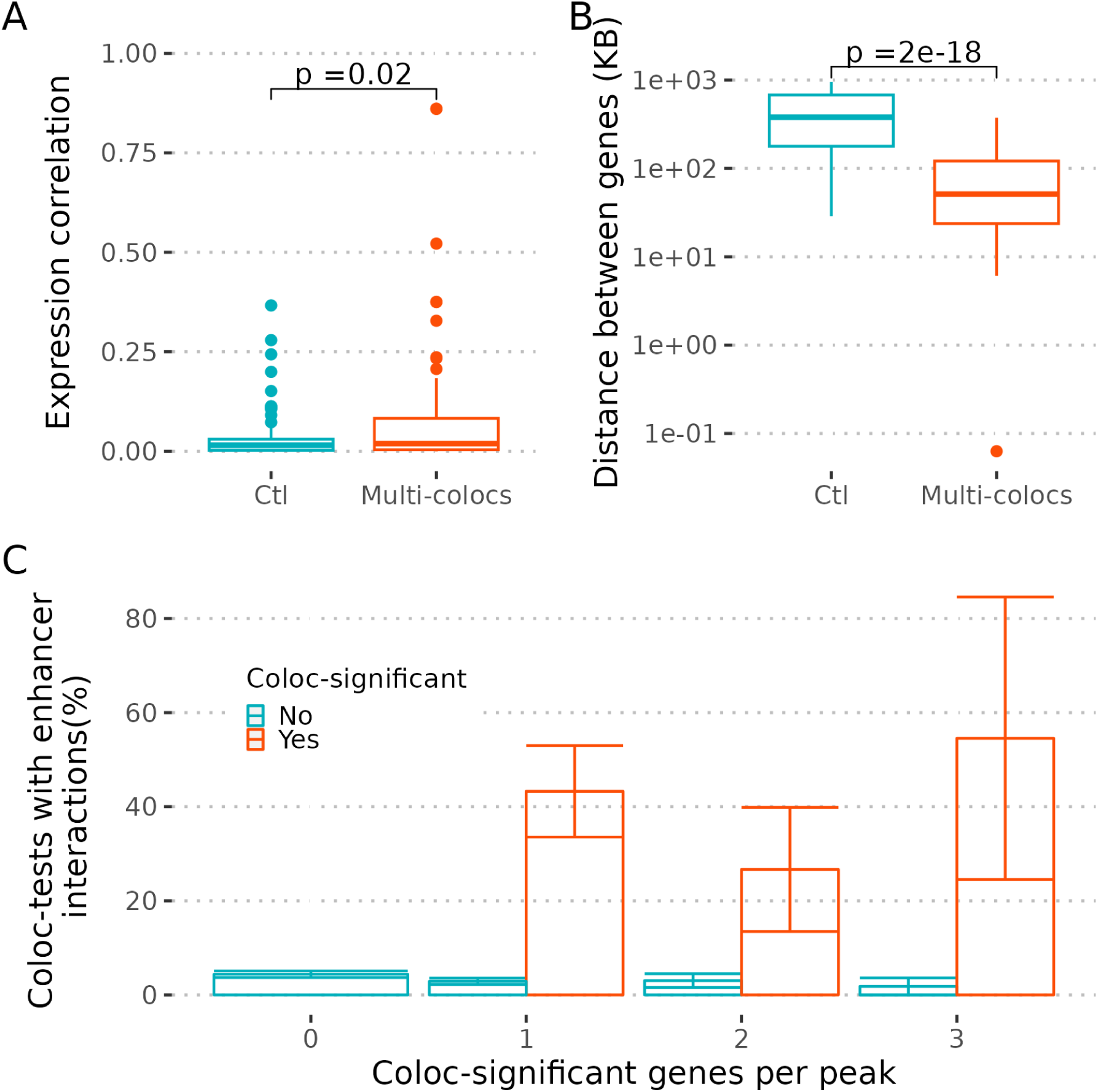
Evidence for co-regulation of multi-coloc genes. **A**. Here, for each eGene we selected the eQTL study with the highest PP(H4) for a given peak. Then, for each pair of significant coloc eGenes a control pair was designed by randomly selecting one of the two coloc eGenes and making a new pair with the selected eGene and a non-significant coloc eGene tested with the same peak and expressed in the same cell type of the non-selected eGene. The non-significant coloc genes were only selected once. This procedure gives the same number of tests and control pairs while matching cell types. The plot shows the distribution of the squared expression correlation (y-axis) for multi-gene colocalisations and controls (x=axis). The p-value for the mean difference is indicated. **B**. Using the same groups as in A, the distance between the TSS of the genes in a pair was calculated and the distribution is shown. The p-value for the mean difference is indicated. **C.** We selected GWAS peaks with the lead variant overlapping an enhancer in immune cells (21%). eQTL studies were aggregated by cell types matching the cell types with enhancer annotations (monocytes, dendritic cells, CD4 T, CD8 T, NK and B cells). Then, for genes with multiple coloc tests within the aggregated cell types, we selected the coloc test with the maximum PP(H4). Next, we stratified peaks by the number of coloc significant genes (0-3). For the peaks with 1 or more significant coloc genes, we stratified the coloc tests by significance (blue/orange). Last, for each group of coloc tests we calculated the proportion of predicted enhancer interactions with the gene promoters matching the cell type of the eGene and the enhancer, which is shown in the y-axis.

Last, we tested whether colocalising genes could be independently linked to their colocalising GWAS peaks, where those GWAS peaks overlapped an enhancer, based on the publically available maps for enhancer-gene interactions in immune cells using the scE2G model^10^ (methods). We found that 40% of GWAS lead SNPs with at least one colocalising gene were located in enhancers linked to their coloc significant eGenes, compared to 3% for GWAS lead SNP/eGene pairs which did not colocalise (p=10^-10^). This proportion did not differ significantly when we stratified peaks by the number of colocalised genes (p=0.9, Fig 5C). A number of GWAS peaks overlapped active enhancers that colocalised with expression of two or more nearby genes. For example, a multiple sclerosis (MS)–associated peak on chromosome 19 lies proximal to *CD37* and *DKKL1*. The overlapping enhancer is predicted to contact the promoters of both genes in B cells, and colocalisation analysis indicates that the MS signal colocalises with cis-eQTLs for *CD37* and *DKKL1* in B cells (Fig. 6A). The two genes show modestly correlated expression in B cells (Pearson *r* = 0.20, *p* = 0.001). Functionally, *CD37* is implicated in B-cell survival and proliferation, whereas *DKKL1,* expressed at low levels in B cells, is principally known to regulate Wnt signalling, and shows high expression in testis. Although both genes are linked to the same regulatory region, current evidence does not establish whether both influence MS risk; it remains plausible that only one is causal while the other is a correlated target of the enhancer.

**Figure 6.**
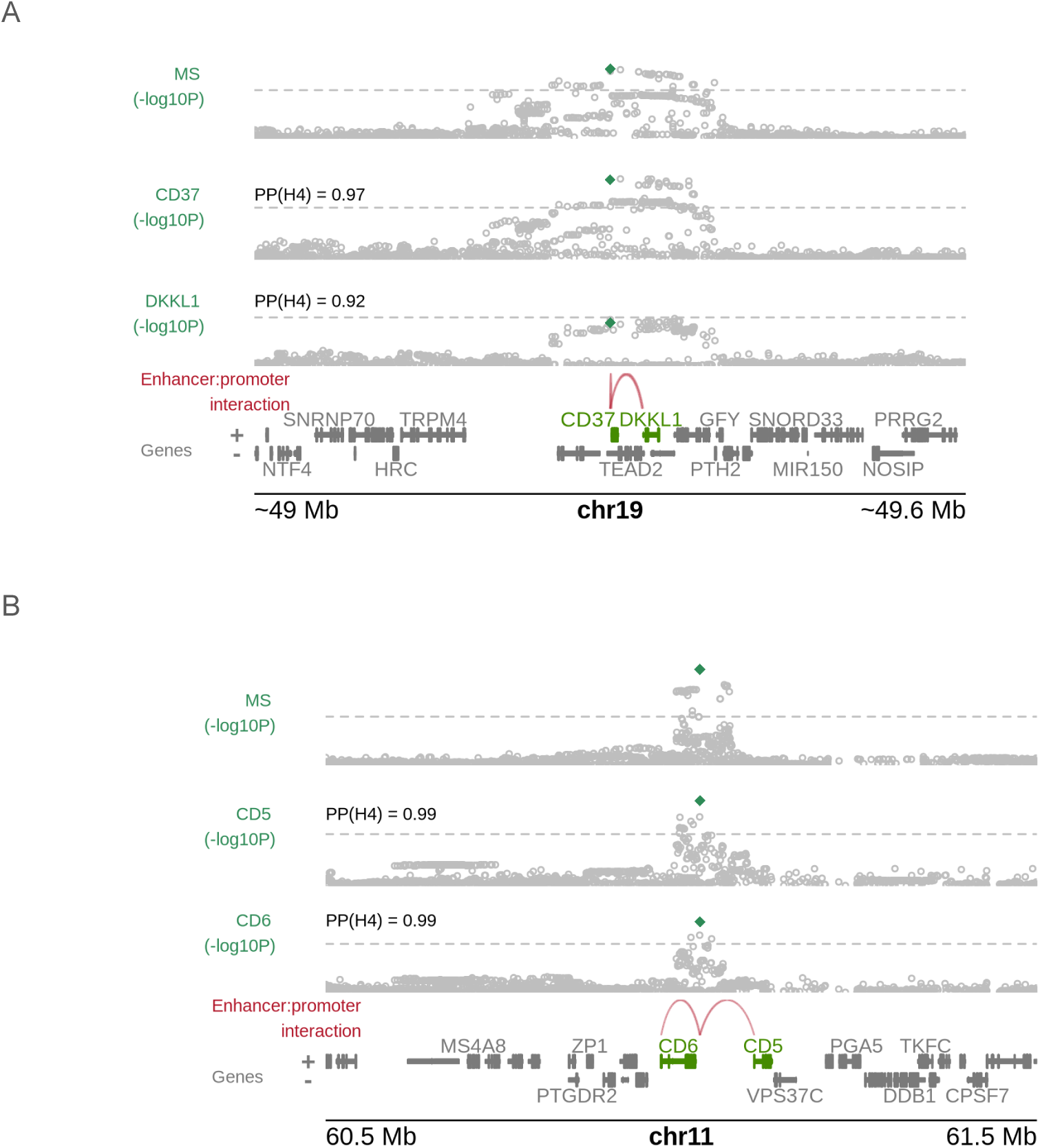
Examples of multi-coloc genes. **A**. Genomic region for MS signal in chromosome 19. The top panel shows the Manhattan plot for MS. The second and third panels show eQTL plots for *CD37* and *DKKL1* respectively from B cells, the posterior probability for the colocalisation hypothesis is indicated (PP(H4)). The dashed grey line corresponds to the genome wide significant level (p=5x10-8). The green symbol corresponds to the lead GWAS SNP. The interactions between the enhancer overlapping the lead GWAS and the gene promoters is indicated by the red arcs. The genes in the region are depicted in the figure, with *CD37* and *DKKL1* in green. **B.** Same as A but for the MS peak in chromosome 11 that colocalised with *CD5* and *CD6* in CD8 T cells.

A second example is an MS-associated peak on chromosome 11 near the paralogous genes *CD5* and *CD6*. The enhancer overlapping this peak is predicted to interact with both promoters in CD8, ⁺ T cells, and eQTL colocalisation provides strong support that the GWAS signal and expression of *CD5* and *CD6* share a causal variant in CD8⁺ T cells (Fig. 6B). The genes are highly co-expressed in CD8⁺ T cells (Pearson *r* = 0.70, *p* < 10⁻⁶), consistent with coordinated regulation by shared regulatory elements; both genes function in T-cell receptor signalling. Although it is currently unknown whether these genes influence MS risk, given the shared functionality, it may be possible that both are causally related.

## Discussion

Although colocalisation was originally devised to be deployed in a semi-hypothesis driven setting, integrating lipid trait GWAS with liver eQTLs^20^, it is now often deployed in a massively parallel approach, taking all available molecular QTL studies compared to one or many GWAS. In this study, we set out to characterise the results of massively parallel colocalisation, with a goal to make recommendations as to the optimal design of colocalisation studies. Granularity and sample size were inversely correlated in the eQTL studies we considered, with a concomitant increase in the number of eQTLs detected in lower-granularity studies. Despite this, eQTLs in higher-granularity studies were more likely to colocalise, which tallies with the cell-specificity of colocalisations. On the other hand, the large sample size in the lowest granularity study - whole blood - enabled the largest total number of colocalisations to be detected.

Therefore, filling the colocalisation gap will require a variety of approaches. One direction is to extend the molecular QTL catalogue to include protein splicing and eRNAs. Another is to continue to expand the breadth of cells captured by high granularity eQTL studies. However, we may most simply gain power through larger sample sizes in molecular QTL studies^16^. This could be achieved through meta-analysis of eQTL studies, although care needs to be taken about carefully matching cell type definitions to avoid obscuring the cell-specific nature of colocalisations. However, at least one large scale single cell eQTL study is in progress, which will have the potential to provide both high granularity and high sample size resources in peripheral blood mononuclear cells: TenK10K^21^. It is likely that single-cell eQTL colocalisation studies to maximise granularity, complemented by large scale meta analyses of the more common cell types will be needed to fully explore the colocalisation space. We also saw higher colocalisation rates in samples including patients for relevant diseases. Here it is hard to see how either large single cell studies or very large eQTL meta analyses of sorted cells will be achieved, though the more modest single cell studies in a few hundred individuals are likely to be invaluable to access cells from a disease context.

We note that colocalisation with coloc can be run in two modes: either under a single causal variant assumption or allowing for multiple causal variants by fine mapping with SuSiE^22^ first. The former is computationally faster, and avoids the need to carefully match reference LD matrices to each GWAS and eQTL dataset, while the latter is likely to discover more colocalisations. Both studies here used coloc with a single causal variant assumption, therefore our focus in this analysis was the relative detection of colocalisation between different eQTL study designs rather than the absolute number of colocalisations. The new Open Targets Platform (https://platform.opentargets.org/) includes a fine mapping step where possible, which would allow a comparison with single and multiple causal variant approaches in future.

While increasing the number of cell types studied increases the chance of a GWAS signal colocalising with at least one eQTL, we also saw a concomitant increase in the average number of colocalised genes per GWAS signal. With multiple testing, there is always the risk of increasing the number of false positives with increasing tests, which is why we used an FDR-based threshold to declare colocalisation. We note that the thresholds estimated in each of these large studies, 0.86 and 0.88, are higher than those typically used in colocalisation analysis. The optimal threshold will depend on the downstream consequences of “calling” a colocalisation. If there is a need to limit false discoveries, then we would advocate the use of an FDR-based approach to determining a threshold. But if colocalisation is a screening step towards designing a large-scale molecular experiment, then a more liberal threshold may be appropriate to ensure true positives are not missed. As ever, there is no one size fits all, and we encourage practitioners to think carefully about the appropriate threshold for their circumstances.

Our initial concern seeing multiple gene colocalisations was that they may reflect inadequate control of false positives. Indeed, methods have been developed to identify the single causal gene in the presence of multiple colocalisation signals, under the assumption that only one gene can be causal^23^. However, sensitivity analysis suggested that even very stringent thresholds did not substantially lessen the issue in either of the studies we considered. Instead, we observed evidence suggesting co-regulation of these “multi-coloc” genes could be at least part of the explanation. Co-regulation of multi-coloc genes clearly complicates interpretation, because only one of the genes may be on the causal pathway, with the remainder acting effectively as biomarkers for the disease-risk variant, and possibly for the causal mechanism^17^. An example is provided by the MS-associated GWAS peak that colocalises with *CD37* and *DKKL1* in B cells. Although it remains unclear whether either gene is causally linked to MS, their weak expression correlation and apparent involvement in distinct signalling pathways suggest divergent biological functions, making it plausible that only one gene is directly causal. On the other hand, multiple colocalising genes may act in concert, with both contributing to disease pathogenesis. It is possible this pattern underlies the colocalisation of the paralogous genes *CD5* and *CD6*, both key regulators of T-cell receptor signalling, with another MS-associated signal.

In the case of co-regulation, observational studies are insufficient to disentangle causality. Experimental perturbations, for example using CRISPR-cas9 based approaches combined with molecular readouts will be required to determine which gene(s) mediate the effect. One possible strategy to guide these experiments is to construct transcriptional networks from healthy and patient derived samples to infer whether the signalling pathways regulated by the multi-coloc genes converge in a downstream process and what differences can be inferred between patients and controls. Such network analysis could guide the selection of molecular targets and readouts for experimental perturbation at the cellular level. Finally, our analysis also reveals that a considerable number of non-coding RNAs colocalised with disease associated variants, highlighting that we remain in the early stages of functionally characterising these regulatory transcripts and their potential contribution to immune-mediated disease.

Another consideration in this new paradigm, where more than one gene frequently colocalises with a single GWAS peak (this study, GTEx^1^ and OTG^3^), is that conclusions for prioritising genes for functional follow-up studies may vary depending on the sources and scope of molecular QTL data used. For instance, in our study, we observed that an MS signal in chromosome 19 colocalised both with *CD37* and *DKKL1* in B cells. A previous study nominated *DKKL1* as the putative causal gene based on colocalisation with a pQTL^24^ ; however, that platform did not include CD37. In contrast, yet another study prioritised *TEAD2* for the same MS locus by integrating eQTL data with promoter capture Hi-C (PCHiC), which identified an open chromatin region in CD4⁺ T and B cells overlapping the lead GWAS variant and interacting with the *TEAD2* promoter^25^. In this case, the GWAS peak SNP was too close to the CD37 transcription start site (∼7 kb) to assess promoter interaction. Note that we also see colocalisation with *TEAD2*, but only in whole blood. This example underscores the importance of integrating multiple data types while carefully considering biases and data gaps that may influence causal gene prioritisation. We suggest that triangulation of evidence is going to become increasingly important to prioritise causal genes as the breadth of molecular data continues to grow.

## Materials and Methods

### GWAS datasets

We collected 22 publicly available, well-powered GWAS datasets (minimum N cases = 5,201), including eight immune-mediated diseases (IMD): autoimmune thyroid disease [AITD]^26^ atopic dermatitis [ATD]^27^ asthma^28^, inflammatory bowel disease [IBD]^29^ multiple sclerosis [MS]^30^ rheumatoid arthritis [RA]^31^ systemic lupus erythematosus [SLE]^32^ and type 1 diabetes [T1D^28,33^].This included two asthma subtypes (child-onset [Asthma CO] and adult-onset [Asthma AO])^34^ and two IBD subtypes (Crohn’s disease [CD] and Ulcerative colitis [UC])^29^ To compare discoveries in immune and non-immune-mediated traits, we included three psychiatric/neurological disorders (Alzheimer’s^35^ major depression disorder [MDD]^36^ and schizophrenia [SCZ^37^]), and three non-immune-mediated conditions (myocardial infarction [MyoInf]^28,38^ osteoarthritis [OArth]^39,40^ and type 2 diabetes [T2D^28,33^]). The ethnicity of the GWAS participants is described in Supp. Table 2.

All datasets were processed using an in-house pipeline to ensure they all have GRCh38/hg38 coordinates and conform to a standard format with minimum requirements of allele, effect size, standard error, and p-value information for all SNPs (see github.com/GRealesM/GWAS_tools).

### eQTL datasets

We collected 101 publicly available blood-cell eQTL datasets from 12 studies^1,11–15,41–46^ and 13 primary cell types: B cells, CD4+ T cells, CD8+ T cells, double-negative T cells (dnT), gamma-delta T cells (gdT), mucosal-associated invariant T (MAIT) cells, NK cells, dendritic cells (DC), hematopoietic stem and progenitor Cells (HSPCs), monocytes, neutrophils, platelets, and whole blood. These datasets include bulk- and single-cell RNA-seq experiments from healthy volunteers and IMD patients of European, African, and Japanese ancestry, with sample sizes ranging from 82 to over 30,000. We also incorporated datasets of naive cells from healthy volunteers stimulated with viral and bacterial stimuli (Supp. Table 1).

Most datasets were obtained from the eQTLcatalog^47^ and required minimal pre-processing. For the eQTLGen whole blood dataset^11^, we used the UCSC liftOver tool to get the GRCh38/hg38 coordinates. For the ImmunexUT datasets^12^ we used data from the Japanese (JPT) sample of 1000Genomes Project Phase III to estimate the allele frequencies. To allow cross-study comparisons we normalised effect sizes (β) and corresponding standard errors (SE) by the estimated standard deviation of the trait 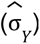. Allele frequencies were used to 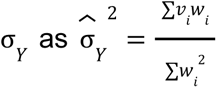 where 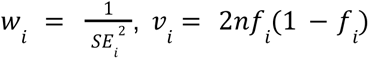 and *n* is sample size, *f_i_* is MAF of SNP *i* and SE_i_ is the standard error of the effect estimate, β*i* at SNP *i*. From these, we calculate corrected estimates and standard errors 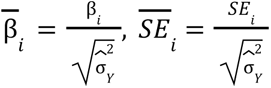. To identify eGenes for testing, we took the lowest p-value in each gene, applied Bonferroni correction (ie. lowest p-value × number of SNPs in that gene), and applied the FDR procedure to the resulting Bonferroni-adjusted p-values, selecting the genes with FDR < 1%.

### OTG colocalisation datasets

We downloaded data from Open Targets Genetics, files now archived at https://ftp.ebi.ac.uk/pub/databases/opentargets/genetics/latest/v2d/.

### Colocalisation in IMDs and QC

We applied a colocalisation and QC procedure on each eGenes across all eQTL-GWAS combinations (22 GWAS and 101 eQTL datasets). For each combination, we selected GWAS regions overlapping eQTL gene regions (typically 2Mb-long) with at least one significant (p < 5 x 10^-8^) SNP. We used the statistical package *coloc*^48^ to assess the evidence of colocalisation. Coloc requires that input SNPs are present in both datasets, producing unreliable results if the strongest associated SNPs for one dataset are missing in the other. To control for this, we used the finemap.abf() function to compute single causal variant finemap posterior probabilities 𝑝_1𝑖_, 𝑝_2𝑖_ for traits 1 and 2, and SNPs *i*. We calculated cross-trait summed posterior probabilities of shared SNPs *S* as 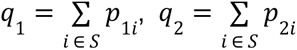 proceeding only if 𝑚𝑖𝑛(𝑞_1_, 𝑞_2_) >. 9, to ensure the inclusion of the most likely causal SNPs.

This resulted in 221,993 coloc tests performed. We ran *coloc* under a single causal variant, with a conservative prior for the hypothesis of colocalisation to reduce the chance of false positive calls (p12 = 5e-6)^49^ . To minimise spurious results, we discarded tests associated with deprecated genes in Ensembl (version 112).

### Significance threshold for colocalisation

We called significant tests those with FDR < 0.05, both in OTG and our in-house analysis. For each test, coloc outputs the posterior probability of colocalisation, PP(H4). We estimated the FDR as the mean of (1-PP(H4)) among the tests with PP(H4) > α and selected α as the largest value such that estimated FDR < 0.05.

### Definition of independent GWAS signals (peaks)

While coloc is performed on regions (typically 2 mb surrounding a transcriptional start site), it is helpful to annotate colocalisations with the GWAS peak. While a test region may contain multiple causal variants, under single causal variant mode coloc will tend to identify colocalisation if any GWAS causal variants colocalise with any eQTL causal variant. To annotate the colocalisation with a GWAS peak, we downloaded fine-mapping 95% credible set data from the OpenTargets platform for each GWAS dataset on 30/5/2025. We used the lead SNPs in each credible set as a representative of independent GWAS signals (henceforth named peaks) to link our coloc tests to for downstream analyses. For each test eQTL region, we checked if it contained any GWAS peak for its corresponding GWAS dataset, assigning the peak to the test if only one was found. For 29,472 regions (including 277 coloc-positive) which contained more than one peak, we visually inspected the multiple peak location in the context of GWAS and eQTL Manhattan plots for all coloc-positive and a sample of coloc-negative tests. Upon inspection, we decided either to merge peaks into one signal when they were considered too close to reliably tell which OpenTargets peak was driving the colocalisation result (retaining the one with the smallest p-value), or manually assigned the coloc test to one of the peaks in the region when this could be clearly distinguished. For regions containing only tests with no evidence of colocalisation and with more than one peak, we merged peaks < 30 kb apart. The 30kb threshold was chosen from examination of the distance between merged/unmerged peaks in visually inspected cases. When no merge happened, we let coloc-negative tests be assigned to multiple peaks, since coloc had rejected colocalisation with either peak. The number of peaks and colocalisation tests assigned by GWAS dataset are summarised in Supp Table 2.

### Determinants of colocalisation

We used the top GWAS peak SNP as a proxy for the causal variant and ran a logistic regression to estimate the log(odds) of colocalisation in any test performed. For the analysis using the OTG colocalisation results the independent variables were the p-value of the GWAS lead SNP, the European MAF and the distance in kilo base-pairs from the lead SNP to the proximal transcription start site (TSS). For the in-house colocalisation analysis of IMDs, an indicator variable was included to capture whether the lead SNP overlapped a predicted active enhancer in immune cells^10^.

### Colocalisation enrichment analysis in OTG

We tested whether cell types or tissues (37) were enriched for colocalisation for immune mediated diseases (3,032 coloc tests), endocrine system disease (8,136 coloc tests) and cardiovascular disease (29,265 coloc tests). For each disease group we ran a logistic regression with significant colocalisation as outcome and independent indicator variables for each cell type or tissue and the total number of tests per cell type or tissue. To account for the clustering structure with multiple observations per GWAS peak, cluster-robust standard errors were computed using the `vcovCL()` function from the *sandwich* package in R. The resulting p-values were Bonferroni-adjusted by the number of cell types or tissues tested. The reference tissue was “adipose”.

### Colocalisation enrichment analysis in IMDs

We grouped colocalisation tests by nominal cell types: B cells, CD4 T cells, CD8 T cells, DC, monocytes and natural killer or whole blood. We estimated the log(odds) of colocalisation using a logistic regression model with cell type as independent variable and a covariate for the number of colocalisation tests per cell type. We use whole blood as the reference category. To assess the enrichment of granularity and sample composition covariates, we excluded whole blood, and re-ran the logistic regression with cell type, sorted vs single cell, mixture of patients/control samples vs control, resting vs stimulated cells as independent variables. We use natural killer cells, sorted cells, controls and unstimulated cells as baseline categories. To account for the clustering structure with multiple observations per GWAS peak, cluster-robust standard errors were computed using the `vcovCL()` function from the *sandwich* package in R. Similar analysis was performed to estimate the log(odds) for colocalisation by nominal cell type for each IMD. Only IMDs with at least 10 coloc tests were considered.

### Co-expression of multi-coloc genes in the same cell type

For this analysis we excluded colocalisation tests in whole blood. We grouped coloc test in the following cell types: B cells, CD4 T cells, CD8 T cells, dendritic cells, monocytes, neutrophils, natural killer cells, gamma-delta T cells, MAIT, double negative T cells and platelets. To avoid redundancy of coloc tests due to overlapping peaks from different IMDs testing the same eQTLs, we only keep one peak randomly, for those with the top fine-mapped SNP within 30KB. We chose 30KB to match our definition of independent peaks (see Definition of GWAS peaks section). This reduced the dataset to 549 peaks. For each gene-gene pair combination tested for colocalisation with the same peak, we annotated whether the two genes significantly colocalised and whether they were expressed in the same cell type. A logistic regression model was employed to estimate the odds ratio for co-localisation among genes expressed within the same cell type. To account for the clustering structure inherent to each peak, cluster-robust standard errors were computed using the `vcovCL()` function from the *sandwich* package in R.

### Expression correlation of multi-coloc genes

For this analysis we selected peaks with two or more significant colocalisation genes in the ImmunextUT dataset which has available gene expression data, resulting in 53 peaks. For each gene-peak pair, we selected the coloc test (performed in different cell types) with the highest PP(H4), resulting in 1175 coloc tests of which 132 were significant. This is to avoid dealing with multiple potentially redundant tests when a significant coloc gene is detected in multiple studies. By selecting the eQTL study with the highest pH4, we ensure we have a good number of tests. Next, for each peak we tabled all pair-wise combinations of significant coloc genes and calculated the gene expression (log +1) Pearson-correlation squared matching the cell types of the coloc tests. As control, for each pair of significant genes, we randomly replaced one of the genes with a non-significant gene tested for the same peak in the same eQTL study as the gene to replace, and we calculated the expression correlation as before. Of note, non-significant genes used as controls were only selected once. This resulted in the same number of correlations for significant coloc tests and controls. A linear regression model with the outcome variable the squared expression correlation of gene pairs and independent variable colocalisation status (significant vs. non-significant) was fit. To account for the clustering structure inherent to each peak, cluster-robust standard errors were computed using the `vcovCL()` function from the *sandwich* package in R.

In addition, we evaluated whether pairs of co-colocalised genes tend to be more proximal than control pairs. We fit a linear model with outcome variable the distance between the transcription start site of the genes in base pairs and explanatory variable whether the pair of genes co-colocalised with the same peak or not. Here, we also computed cluster-robust standard errors using the `vcovCL()` function from the *sandwich* package in R to account for the multiple colocalisation tests per peak.

### Testing whether GWAS peak-lead SNPs tend to overlap regulatory elements for coloc eGenes

We used publically available maps for enhancer-gene and promoter-gene regulatory interactions using the scE2G multiome model for dendritic cells, monocytes, B cells, CD4 and CD8 T cells and natural killer cells from human peripheral blood mononuclear cells^10^. The input for the scE2G multiome model is paired scRNA and scATAC-seq data, and using a classifier based on logistic regression the model predicts which chromatin accessible elements act as enhancers to regulate which genes in a given cell population. Each enhancer-gene pair is annotated with a score (E2G score) corresponding to the probability of a regulatory effect. Active enhancer-gene pairs were defined as those with an E2G score > 0.164, as previously described^10^. Subsequently, we selected GWAS peaks for which the lead SNP overlapped a predicted active enhancer in any of the immune cell types tested (monocytes, dendritic cells, CD4 T, CD8 T, NK and B cells), under the assumption that the causal variant regulates the enhancer activity. We found that ∼21% of the peaks overlapped an active enhancer. Then, for genes that had more than one coloc test with a peak within a nominal cell type (monocytes, dendritic cells, CD4 T, CD8 T, NK and B cells) we selected the coloc test with the maximum PP(H4). A mixed-effects logistic regression model was fitted using the `glmer()` function from the *lme4* package in R. The binary outcome variable indicated whether the enhancer overlapping the peak lead SNP interacted with the eGene promoter in the same cell type of the eQTL study. Fixed effects included (i) an indicator variable denoting whether the colocalisation (coloc) test was significant and (ii) the number of coloc significant genes associated with a peak. An interaction term between these two predictors was included to test for effect modification. To account for non-independence arising from multiple coloc tests per peak, a random intercept for peak was incorporated into the model.

## Data availability

Our in-house colocalisation analysis can be found in Supp. Tables 3 and 4.

## Code availability

The code for harmonising GWAS summary statistics can be found at github.com/GRealesM/GWAS_tools.

The code for making the figures can be found at https://gitlab.com/evigorito/coloc_gap

## Acknowledgements

This work was co-funded by the Wellcome Trust (WT220788) and the MRC (MC_UU_ 00002/4).

---

**Supp. Figure 1.**
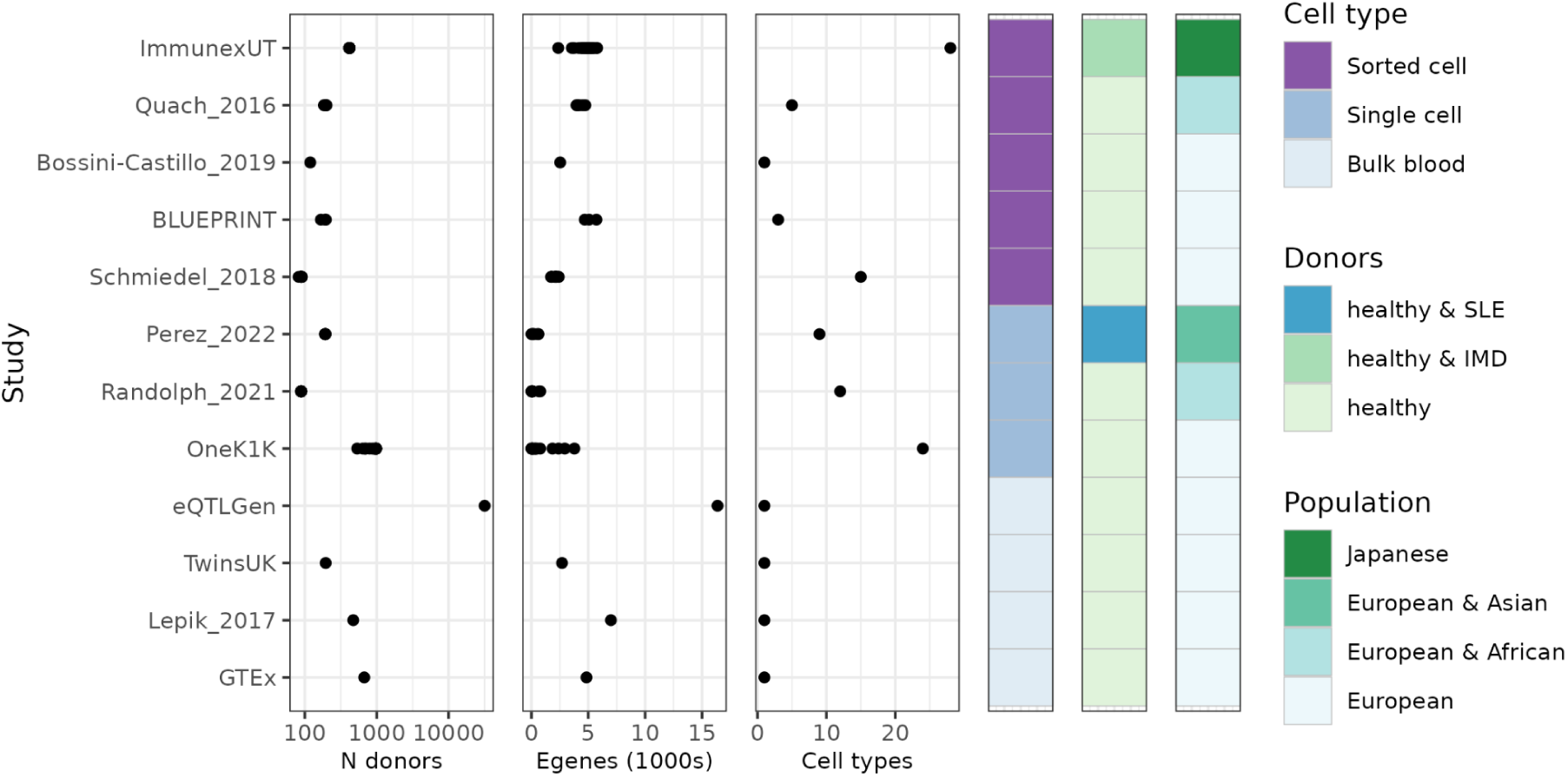
Summary of eQTL studies. Diversity of the study design of the 101 eQTL experiments used in our analysis (x-axis) from 12 studies (y-axis).

**Supp. Figure 2.**
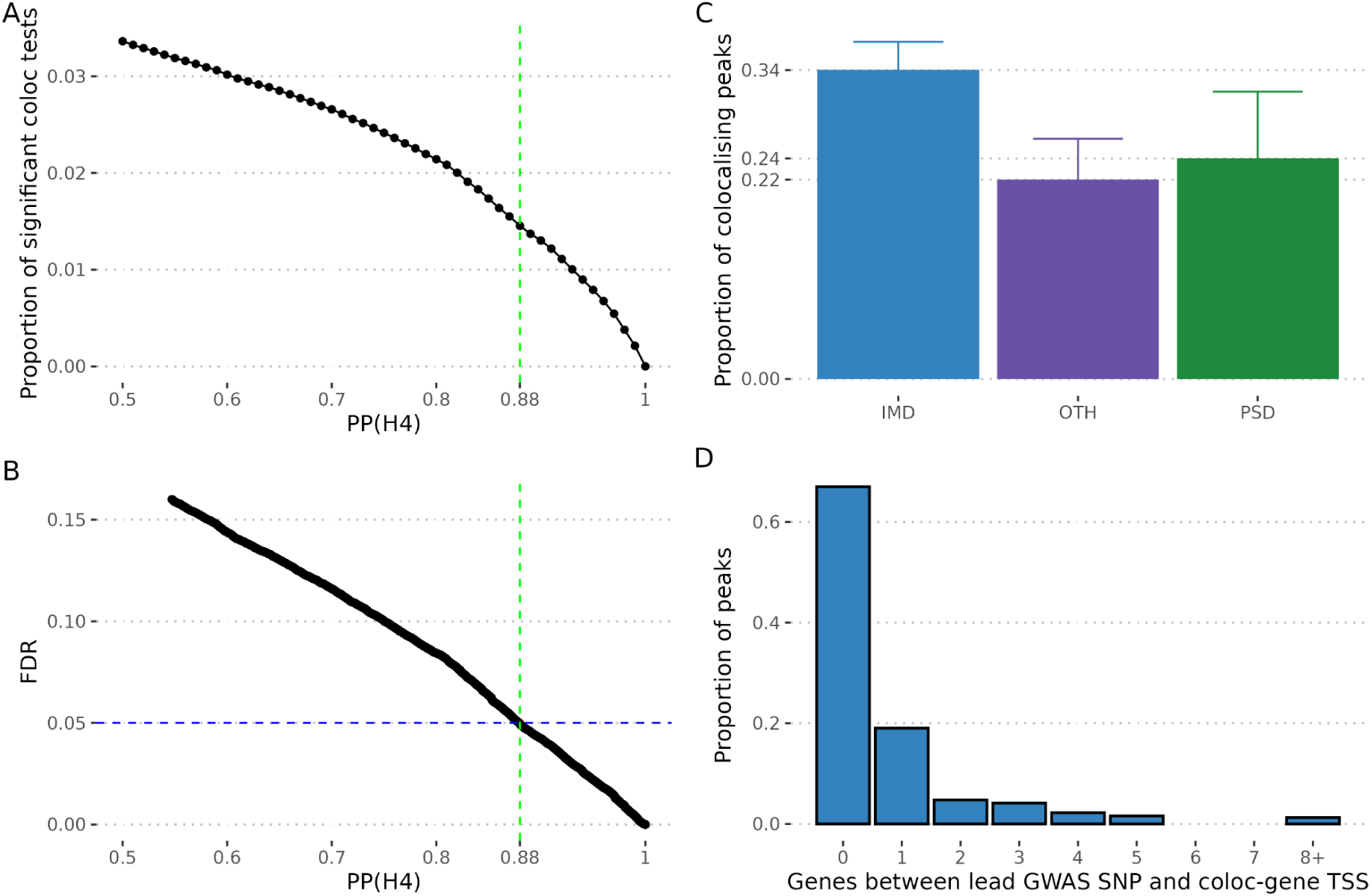
Colocalisation in IMDs and related diseases. **A.** Proportion of coloc significant tests (y-axis) as a function of the posterior probability of colocalisation (PP(H4)). **B.** Estimated false discovery rate (FDR) (y-axis) as a function of the posterior probability for colocalisation (PP(H4)) (x-axis). The vertical and horizontal lines indicate the threshold selected from downstream analysis that corresponds to 5% FDR. **C**. Proportion of peaks with evidence of colocalisation comparing IMDs (blue bar), diseases with underlying chronic inflammation but not considered immune mediated, OTH (purple bar) or psychiatric disorders, PSD (green bar). The error bars correspond to the 95% confidence interval. **D.** Number of gene TSS between an IMD GWAS lead SNP and its closest significant coloc eGene (truncated at 8).

**Supp. Figure 3:**
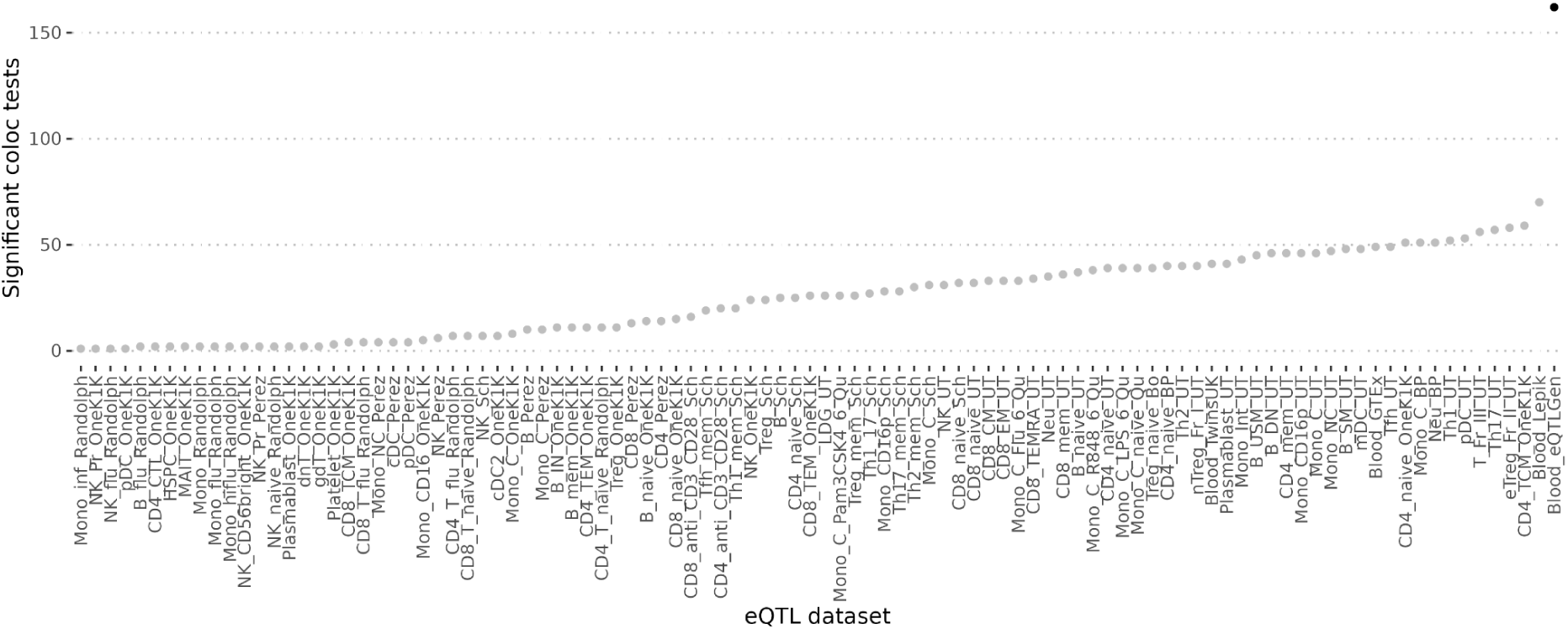
Number of significant colocalisations by eQTL study in IMDs. For each of the eQTL studies (x-axis) the dots show the absolute number of coloc significant tests across all IMDs (y-axis). eQTLGen is shown in black.

